# Quality of life and patient-reported outcomes following proton therapy for oropharyngeal carcinoma: a systematic review

**DOI:** 10.1101/2020.11.04.20226175

**Authors:** Noorazrul Yahya, Hanani A. Manan

## Abstract

**Background:** Complex anatomy surrounding the oropharynx makes proton therapy (PT) especially intensity-modulated PT (IMPT) a good option due to its ability to reduce the volume of irradiated healthy tissues. Dosimetric improvement may not translate to clinically relevant benefit. As outcome data are emerging, we aimed to systematically review current evidence of the quality of life (QOL) and patient-reported outcomes (PRO) following PT of oropharyngeal carcinoma (OC).

**Materials and Methods:** We searched PubMed and Scopus electronic databases to identify eligible reports on QOL and PRO following PT of OC according to PRISMA guidelines. Reports were extracted for information on demographics, main results and clinical and dose factors correlates.

**Results:** Six reports were selected from which four compared between PT and photon-based therapy. All endpoints where significant differences were found favour PT. The change following treatment improves but never to the baseline level.

**Conclusion:** Available evidence suggests that PT causes less QOL deterioration and PRO compared to photon therapy. Whether or not it is cost-effective should be a subject of further investigation.

## INTRODUCTION

Among patients treated for oropharyngeal carcinoma (OC), acute and late treatment-related sequelae including xerostomia, dysphagia and dysgeusia remain to be a challenge [1-3]. These consequences of treatment may impact the quality of life (QOL) including difficulties in communication, nutritional intake enjoyment and social contact [2]. Maintaining good QOL following OC is especially important now due to improved survivorship related to better treatment regime and improved prevalence of human papillomavirus (HPV)-related squamous cell carcinoma which disproportionately impacts young and physically fit individuals [4]. Thus, improving therapies to mitigate the occurrence of these treatment side effects is paramount.

As there are multitudes of studies which have empirically demonstrated the impact of dose distributions including to salivary glands [5-10], swallowing muscles [8-12], oral cavity [2] and other normal tissues to patient-reported outcomes (PRO) and quality of life (QOL) measures in photon-based therapy. Proton therapy (PT) offers several advantages over photon therapy in including the elimination of exit dose and the possibility to tailor the dose resulting from the physical characteristics of the beams with Bragg peaks [13, 14]. Several dose comparison studies have shown significant dose reduction favouring PT [15, 16]

While dosimetric analyses have proven the benefits of PT based on dosimetric advantage [17, 18], PT is not without its challenges [19]. Deviation of the given dose distribution from the intended distribution due to uncertainties from intrafraction motion and patient set-up error is especially larger in pencil beam scanning technique. Furthermore, range uncertainties as a systematic error can be in the range between 3 to 3.5%. Combining these two sources of errors result in the radiological path length of proton beams which are different from the intended length. While the simulation of radiation is highly accurate, the modelling of the human body based on CT images of the patient is not similarly precise due to the calibration uncertainties between the Hounsfield unit (HU) values and the proton stopping powers and intra- and interfractional variations in anatomy [20]. Beddok et al. in a review has outlined the challenges and how they can be at least partially overcome [20]. These errors are relevant to normal tissue effects as improved dose distribution may not necessarily improve PRO and QOL measures.

There appear to be a few reports presenting the actual impact of PT to PRO and QOL. The limited availability of PT in the past may be the reason for the sparse empirical evidence. Furthermore, some institutions prefer to assess toxicities based on physician-reported measures probably due to the historical emphasis on these measures. However, as more centres are offering PT and increasing interest to measure outcomes which are more patient-centred and connect to patients in a more meaningful way, we may see more clinical results especially involving PRO and QOL mature. In this study, we aimed to systematically review the PRO and QOL changes following PT of oropharyngeal carcinoma.

## MATERIALS AND METHODS

### Systematic review protocol and eligibility criteria

The systematic review protocol and methodology established by Preferred Reporting Items for Systematic Reviews and Meta-Analyses (PRISMA) was utilised [21-23]. Original research manuscripts were evaluated for inclusion or exclusion based on PICOS criteria detailed in Supplementary A. The PICOS framework is used to develop literature search strategies by systematically determine the inclusion based on patient population, intervention, comparison, outcome, and study design. Reports fulfilling all five criteria were included. Excluded studies were reported based on the first PICOS criterion not fulfilled.

### Search strategy and selection process

Electronic databases (National Center for Biotechnology Information (PubMed) and Scopus) were searched to identify articles. Keywords used are detailed in Supplementary B. In the first phase, articles were reviewed in increasing specificity via the title, abstract, then finally, via full text by NY and HAM independently. In the second phase, bibliographic references, and citations of relevant studies in phase one were extracted from Scopus and hand searched for additional eligible studies based on the assumption that relevant studies cited or were cited by other related studies. We have confidence in the robustness of this two-step method to ensure no omission of relevant studies. No publication date or publication status restriction was imposed. Discrepancies in the results of the selection were deliberated in team meetings. Where more than one reports of a study existed, reports with a more complete result were included. Where an institution published multiple reports from the same patient cohort but with different endpoints, all reports were included. Study search and selection were completed in September 2020.

### Quality assessment

We used an assessment tool from the National Heart, Lung and Blood Institute; Quality Assessment of Case-Control Studies to evaluate the quality of studies where comparisons were made to patients treated with photon therapy and Quality Assessment Tool for Observational Cohort and Cross-Sectional Studies to evaluate studies comparing outcomes to baseline measures.

### Data review and extraction

Upon finalisation of article selection, data extraction was performed by NY. Information was extracted into spreadsheets and included details of the articles, patients, proton therapy dose regime and technique and measures for QOL or/and PRO. If comparisons were made to photon-based therapy, the treatment and patient characteristics for photon therapy were also extracted.

### Meta-analyses

Due to the small number of eligible studies and studies from the same pool of patients reported by the same groups in multiple articles for different endpoints, meta-analysis is not warranted.

## RESULTS

### Study selection and quality assessment

The database queries produced 80 and 150 records from PubMed and Scopus, respectively (Fig. 1, Supplementary A). After removal of duplicates, 175 reports were reviewed for inclusion and 6 met the inclusion criteria [24-29]. In the second phase, where citations of the previously selected reports were reviewed using Scopus which is a source-neutral abstract and citation database, 240 articles were reviewed, and no additional papers were found. The included studies were found to be of reasonable quality with patients accrued in a centre which is expected due to the limited number of proton centres. Sample size and calculations were rarely mentioned.

**Figure 1:** Identification of inclusion based on PRISMA. Eligibility was determined using PICOS criteria (Supplementary A).

### Characteristics of included studies

Table 2 summarises the characteristics of the selected studies including 302 patients treated with proton and 438 patients treated with photon. Due to the possible overlap of patients across studies reported by the same research groups, the number of patients is likely to be an overestimation and we refrained from summing the number of patients in the preceding subsections. All studies reported prospectively collected outcome measurements. The publication dates ranged from 2016 to 2020 which reflect the recency of PT introductory into widespread practice and emphasis to PRO and QOL.

**Table 1:** Study characteristics

| Reference | No. | Stage III/IV (%) | % HPV positive (+), unknown (?) | Type of proton therapy | Dose (Gy RBE) | Other treatments | Therapy comparison | Details of therapy comparison | QOL measures |
| --- | --- | --- | --- | --- | --- | --- | --- | --- | --- |
| Bagley 2020 | 69 | AJCC7 stage III-IV, M0 - 100 | + 84<br>? 14 |  | Median - 69.3, range 60–70 | Induction -5, concurrent – 38, Induction + concurrent – 11 | - | - | 15-item Xero: Relat Scale (XeQ |
| Blanchard 2016 | 50 | T3-T4 – 20<br>N2-N3 - 80 | + 88<br>? 10 |  | Small volume disease – 66, advanced disease - 70 | Concurrent – 64% | 100 IMRT | 2:1, matched laterality, site, HPV, T and N status, smoking & chemotherapy | Not s |
| Grant 2020 | 71 | AJCC 7th edition stage III/IV - 100 | + 85.9<br>? 7.0 |  | range - 66-70 Gy | Induction -5, concurrent – 41, induction + concurrent – 10, | - | - | MD / Dysp Inver |
| Manzar 2020 | 46 | AJCC 7th edition Stage III/IV – 84.8 | + 76.1<br>? 13.0 |  | adjuvant, range - 60–66; definitive – 70 | Concurrent - 36 | 259 VMAT | Significant difference: age (IMPT older) smoking status and pack-years (VMAT higher), dose category (more definitive RT in IMPT), | EOR: H&N: |
| Sharma 2018 | 31 | Stage I-III - 13 IVA | Not stated |  | median - 61.7 | Chemotherapy - 12 | 33 VMAT | Median dose - 62.6 (ns); chemotherapy | QLQ: versi EOR: |
|  |  | - 87 |  |  |  |  |  | - 14 (ns) | H&N:<br>the G<br>(GRI<br>Xero:<br>Work<br>and<br>Perfc<br>Statu<br>Heac<br>Neck<br>ques |
| Sio 2016 | 35 | Stage<br>III-IV –<br>94.3 | +74.3<br>? 20 |  | median<br>70.0,<br>range -<br>59.0-70.0 | concurrent<br>chemotherapy<br>– all, induction<br>- 26 | 46 IMRT | Significant<br>difference:<br>location (more<br>tonsil), T-stage<br>(more T3-T4),<br>lower<br>induction<br>chemotherapy,<br>higher total<br>radiation dose | MD /<br>Symp<br>Inver<br>Heac<br>Neck<br>(MD/<br>top 1<br>seve |

**Table 2:** Significant endpoints and significant factors impacting the endpoints

| Reference | Significant endpoints | Non-significant endpoints | Significant clinical factors | Do |
| --- | --- | --- | --- | --- |
| Bagley 2020 | General xerostomia including physical, personal, pain, and social domains | - | Time, baseline XeQoLS score, stage and N status | Un<br>Mu |
| Blanchard 2016 | Xerostomia at 3 months (favours IMPT) | Fatigue and xerostomia at 1 year. | Not studied | No |
| Grant 2020 | Composite score for dysphagia | - | T-stage | No |
| Manzar<br>2020 | Overall: Cough, need for nutritional supplements and dysgeusia (favour IMPT).<br>Subgroup analyses: Adjuvant – Cough, sense of smell/taste and problems with teeth, (favour IMPT) and sexual symptoms (favour IMRT).<br>Definitive - Feeling ill, feeding tube, sense of smell/taste & nutritional supplements (favour IMPT).<br>Bilateral - Cough & nutritional supplements (favour IMPT)<br>Unilateral - dry mouth, sticky saliva & sense of smell/taste (favour IMPT).<br>Concurrent chemotherapy – swallow, nutritional supplements, feeling ill & sense of smell/taste (favour IMPT)<br>RT only - cough sense of smell/taste & problems with teeth (favour IMPT) & swallow (favour IMRT) | EORTC H&N QLQ-35 questions not stated in the previous cell | Not studied | Sig<br>bet<br>As:<br>an: |
| Sharma<br>2018 | 3 months – dental problem<br>6 months – moderate-to-severe dry mouth, xerostomia day, xerostomia night, dental problems, physical function, role function.<br>12 months – H&N pain, xerostomis, moderate-severe dry mouth, role function. | Fatigue, sticky saliva (day general) and global health and timepoints not stated in the previous cell | Not studied | Sig<br>bet<br>As:<br>an: |
| Sio 2016 | Symptom score - subacute phase – food taste and appetite<br>Chronic phase – appetite (all favour IMPT)<br>moderate to severe symptoms – subacute phase – food taste and mucus (favour IMPT)<br>mean of top 5 MDASI higher during subacute phase for IMRT | Other top 11 symptoms - Dry mouth, swallowing/chewing, fatigue, pain, sleep, mouth sores, drowsiness, distress. | Not studied | No |

### Studies comparing proton therapy and photon radiotherapy

Four studies presented the comparison between proton therapy and proton radiotherapy in term of QOL and PRO changes (Table 2). Blanchard et al. 2016 and Sio et al. 2016 compared the outcomes of proton therapy to patients treated with IMRT while more recent reports (Manzar et al. 2020 and Sharma et al. 2018) used patients treated with VMAT. Overall, in all QOL and PRO measures where the differences were significant, patients treated with proton therapy reported better outcomes including lesser xerostomia, lesser cough, lesser need for nutritional supplements, lesser dysgeusia, better food taste, better appetite, less mucous and better general symptoms.

### Effect of time

In this review, we divided the time into several time points (Table 3); acute, subacute, late at <1 year and late at ≥1 year. In comparison to patients treated with photon radiotherapy, patients treated with proton therapy have better outcomes across most time points. Sio et al. is the only report which included the outcome during treatment found no difference between the two treatment modalities. Blanchard et al. found the difference for xerostomia during subacute timepoint only and no difference was found during the late time points. Bagley et al. and Grant et al. found a similar pattern for xerostomia and dysphagia where the worst was during the treatment which improved during subacute and late phases. The scores, however, remained worse than baseline.

**Table 3:** Significant endpoints based on the time points divided into acute, subacute, late at <1 year and late at ≥1 year.

### Effect of dose factors

Only one study performed an analysis of dose-outcome association which found a significant univariate association of oral cavity dose to xerostomia [28]. In multivariate analysis, the feature is no longer significant. Doses to multiple structures were found to receive a lower radiation dose in proton therapy compared to photon therapy reported by Sharma et al. and Manzar et al. which complement findings from other dose comparison studies for head and neck cancers [17, 25, 26, 30, 31].

### Effect of clinical factors

The T and N stages, baseline status and time from the radiotherapy were found to be significant predictors for patient-reported and quality of life outcomes.

## DISCUSSION

We conducted a systematic review to methodically accumulate and synthesize the evidence of patient-reported outcomes and quality of life changes following proton therapy of the oropharyngeal carcinoma. This is an improvement from a systematic review by Verma et al. which combined treatments of many diagnoses treated with proton therapy which provided a good breadth of the issue but not depth [32]. Furthermore, we found five new articles which fulfilled our inclusion criteria as compared to one described in Verma et al. in 2017 [32]. This is expected given the exponential growth of the number of reports due to the recent availability of proton therapy in many centres. Based on this systematic review, we found; 1) studies consistently showed the advantages of proton therapy compared to photon therapy, 2) studies showed a significant decline in functions following proton therapy at the acute stage which will improve and 3) functions do not revert to pre-treatment status.

Six reports were included in this systematic review. However, four of which came from a single centre, MD Anderson Cancer Centre in Houston Texas. Furthermore, all centres reporting the outcomes were from the United States of America. The lack of reports from other centres is likely to be associated with the low number of centres which offer proton therapy for the treatment of oropharyngeal carcinoma. Only 109 proton therapy machines are now operational across the world from which 40 are located in the USA (dirac.iaea.org). To put this into perspective, more than 14000 high energy photon therapy machines are available across the globe. As 38 new centres are now under construction [33], more reports on treatment outcomes following proton therapy may come soon. Second, centres are likely to focus on provider-reported outcomes for toxicity following treatments of oropharyngeal carcinoma which are reported in several studies [34]. As the emphasis on PROs and QOL measures is increasing in recent years due to a paradigm change to increase the involvements of patients in decision-making and to capture outcomes which matter to patients [35], we are optimistic to see reports with larger cohorts from other centres across the globe.

Generally, the patient-reported outcomes and the quality-of-life measures favour proton therapy. This can be simply explained by the better dose distribution using proton therapy due to the beam characteristics as several dose comparison studies have observed the superiority of proton therapy as compared to photon [17, 30, 31]. The dose to the salivary gland and the larynx, structures commonly associated with increased risks of xerostomia and dysphagia, were found to be higher when VMAT or IMRT were utilised [30, 36]. However, the difference was only significant within the first year following treatment. The change for xerostomia was found to be no longer significant after the acute phase [24] and the number of domains remained significantly worse for photon therapy after the acute phase was also reduced [27, 37]. At least three hypotheses can be made from this observation. First, the higher dose received from photon therapy causes delayed recovery associated with a higher volume of tissue receiving low dose radiation, common in IMRT. Second, after the initial stage of quality of life changes following cancer treatment, patients may have adapted to the new norms and thus, reported less bother. Third, it should be noted that none of these studies randomised patients into proton or photon treatment arms. Consequently, bias due to systematic differences between patients treated with proton and photon therapy cannot be discounted and probably impacting future cost-effectiveness analysis [38].

The main drawback of proton therapy is the high cost associated with the construction of a cyclotron and maintenance of the facility. To put the comparison in perspective, the cost of proton therapy is approximately 2.4 times more than photon-based therapy. Frequently, the cost-effectiveness of proton therapy is being questioned [39]. Cost-effectiveness analysis showed that the use of IMPT was only cost-effective for a fraction of younger patients with a high risk of profound reduction of long-term morbidity [39]. To identify patients expected to benefit from proton therapy compared to IMRT, selection based on NTCP models has been implemented [40, 41]. In this strategy, planning comparisons between photon and proton therapy were performed on an individual level. The reduction in multivariable normal tissue complication probability (NTCP) models which numerically describes the relationship between the dose delivered to organs at risk and clinical factors and the predicted risk of radiation-induced side effects were then employed to detect whether PT is clinically advantageous. With this strategy, resources will be optimally utilised for the benefit of patients without straining the healthcare system. This NTCP-based method, however, is subject to model- and dose-related uncertainties [42]. Furthermore, the high cost can be financially problematic for countries with limited resources which further widen the gap of health disparities between countries [43-46]. Especially for treatment like oropharyngeal carcinoma where it is expected that only 0 to 0.4% probabilities that proton therapy was cost-effective for 65- and 55-year old patients [39], its utilisation in low- and middle-income country is very likely to be societally unacceptable. Fortunately, the cost for proton therapy is continuously decreasing with more compact designs allowing space-related costs to be further minimised. The treatment delivery is also improving which may further reduce the dose to normal tissue [47].

We should note some limitations of the systematic review. First, a small number of the available study were conducted in a limited number of centres may limit the applicability of this review. Second, due to the lack of independence across studies, no meta-analyses can be performed. Third, it is acknowledged that selection bias where negative studies are less likely to be published and thus not be searchable may influence the observation.

## CONCLUSION

Proton therapy may improve patient-reported and quality of life outcomes for patients treated for oropharyngeal carcinoma especially in acute and early late timepoints which improve in later time points. This observation can be proven or challenged as more data available as proton therapy becomes more widely available.

## Data Availability

All data is available within this manuscript

## Supplementary Materials

Supplementary A: PICOS criteria for inclusion

Supplementary B: Search strategy using Pubmed and Scopus databases

Supplementary C: Quality check of selected studies

## Abbreviations

PT: Proton therapy
PRISMA: Preferred Reporting Items for Systematic Reviews and Meta-Analyses

