## Supplementary for "Quality of life and patient-reported outcomes following proton therapy for oropharyngeal carcinoma: a systematic review"

Supplementary A: PICOS criteria for inclusion

|  | Criteria |
| --- | --- |
| P - patient | adult human patients with oropharyngeal carcinoma |
| I - intervention | all types of proton therapy |
| C - comparison | Comparison to baseline status or photon therapy |
| O - outcome | Patient-reported or quality of life measures |
| S – type of study | Randomised-controlled trials, cohort (prospective or retrospective). Exclude studies with no statistical comparisons (case study or case series) or reviews. |

Supplementary B**: Search strategy using Pubmed and Scopus databases**

| Database | Search string | Articles found |
| --- | --- | --- |
| **Stage 1** | | |
| Pubmed | ("proton therapy"[MeSH Terms] OR ("proton"[All Fields] AND "therapy"[All Fields]) OR "proton therapy"[All Fields]) AND ("oropharyngeal neoplasms"[MeSH Terms] OR ("oropharyngeal"[All Fields] AND "neoplasms"[All Fields]) OR "oropharyngeal neoplasms"[All Fields] OR ("oropharyngeal"[All Fields] AND "carcinoma"[All Fields]) OR "oropharyngeal carcinoma"[All Fields]) | 80 |
| Scopus | TITLE-ABS-KEY (oropharyngeal OR oropharynx ) AND TITLE-ABS-KEY ( proton AND therapy ) AND TITLE-ABS-KEY ( cancer OR carcinoma ) | 150 |
|  | Unique titles | 175 |
| **Stage 2** | | |
|  | Unique citations from selected articles not found in Stage 1 | 240 |
|  | Included | 0 |

Supplementary material B: Quality check of selected studies

**Quality Assessment of Case-Control Studies**

|  | **Study** | | | | | | | | | | | |
| --- | --- | --- | --- | --- | --- | --- | --- | --- | --- | --- | --- | --- |
|  | **Blanchard 2016** | | | **Sharma 2018** | | | **Manzar 2020** | | | **Sio 2016** | | |
| **Criteria** | **Yes** | **No** | Other (CD, NR, NA)* | **Yes** | **No** | Other (CD, NR, NA)* | **Yes** | **No** | Other (CD, NR, NA)* | **Yes** | **No** | Other (CD, NR, NA)* |
| 1. Was the research question or objective in this paper clearly stated? | / |  |  | / |  |  | / |  |  | / |  |  |
| 2. Was the study population clearly specified and defined? | / |  |  | / |  |  | / |  |  | / |  |  |
| 3. Was the participation rate of eligible persons at least 50%? |  |  | CD |  |  | CD |  |  | CD |  |  | CD |
| 4. Were all the subjects selected or recruited from the same or similar populations (including the same time period)? Were inclusion and exclusion criteria for being in the study prespecified and applied uniformly to all participants? | / |  |  | / |  |  | / |  |  |  | / |  |
| 5. Was a sample size justification, power description, or variance and effect estimates provided? | / |  |  |  | / |  |  | / |  |  |  | / |
| 6. For the analyses in this paper, were the exposure(s) of interest measured prior to the outcome(s) being measured? | / |  |  | / |  |  | / |  |  | / |  |  |
| 7. Was the timeframe sufficient so that one could reasonably expect to see an association between exposure and outcome if it existed? | / |  |  | / |  |  | / |  |  | / |  |  |
| 8. For exposures that can vary in amount or level, did the study examine different levels of the exposure as related to the outcome (e.g., categories of exposure, or exposure measured as continuous variable)? | / |  |  | / |  |  | / |  |  | / |  |  |
| 9. Were the exposure measures (independent variables) clearly defined, valid, reliable, and implemented consistently across all study participants? | / |  |  | / |  |  | / |  |  | / |  |  |
| 10. Was the exposure(s) assessed more than once over time? |  |  | NR |  |  | NR |  |  | NR |  |  | NR |
| 11. Were the outcome measures (dependent variables) clearly defined, valid, reliable, and implemented consistently across all study participants? | / |  |  | / |  |  | / |  |  | / |  |  |
| 12. Were the outcome assessors blinded to the exposure status of participants? |  | / |  |  | / |  |  | / |  | / |  |  |
| 13. Was loss to follow-up after baseline 20% or less? |  |  | CD |  |  | CD |  |  | CD |  |  | CD |
| 14. Were key potential confounding variables measured and adjusted statistically for their impact on the relationship between exposure(s) and outcome(s)? | / |  |  | / |  |  | / |  |  | / |  |  |

### Quality Assessment Tool for Observational Cohort and Cross-Sectional Studies

|  | **Study** | | | | | |
| --- | --- | --- | --- | --- | --- | --- |
|  | **Bagley 2020** | | | **Grant 2020** | | |
| **Criteria** | **Yes** | **No** | Other (CD, NR, NA)* | **Yes** | **No** | Other (CD, NR, NA)* |
| 1. Was the research question or objective in this paper clearly stated? | / |  |  | / |  |  |
| 2. Was the study population clearly specified and defined? | / |  |  | / |  |  |
| 3. Was the participation rate of eligible persons at least 50%? |  |  | CD |  |  | CD |
| 4. Were all the subjects selected or recruited from the same or similar populations (including the same time period)? Were inclusion and exclusion criteria for being in the study prespecified and applied uniformly to all participants? | / |  |  | / |  |  |
| 5. Was a sample size justification, power description, or variance and effect estimates provided? |  | / |  |  | / |  |
| 6. For the analyses in this paper, were the exposure(s) of interest measured prior to the outcome(s) being measured? | / |  |  | / |  |  |
| 7. Was the timeframe sufficient so that one could reasonably expect to see an association between exposure and outcome if it existed? | / |  |  | / |  |  |
| 8. For exposures that can vary in amount or level, did the study examine different levels of the exposure as related to the outcome (e.g., categories of exposure, or exposure measured as continuous variable)? | / |  |  | / |  |  |
| 9. Were the exposure measures (independent variables) clearly defined, valid, reliable, and implemented consistently across all study participants? | / |  |  | / |  |  |
| 10. Was the exposure(s) assessed more than once over time? |  |  | NR |  |  | NR |
| 11. Were the outcome measures (dependent variables) clearly defined, valid, reliable, and implemented consistently across all study participants? |  |  |  |  |  |  |
| 12. Were the outcome assessors blinded to the exposure status of participants? | / |  |  | / |  |  |
| 13. Was loss to follow-up after baseline 20% or less? |  |  | CD |  |  | CD |
| 14. Were key potential confounding variables measured and adjusted statistically for their impact on the relationship between exposure(s) and outcome(s)? | / |  |  | / |  |  |
